# Machine learning approach to dissect the clinical heterogeneity of IBD-associated fatigue

**DOI:** 10.1101/2025.08.14.25333676

**Authors:** Cher S Chuah, Rebecca Hall, Robert J Whelan, Peter D Cartlidge, Beatriz Gros, Eva Iglesias-Flores, Nikita Parkash, Ray K Boyapati, Clara Ramos-Belinchon, Solomon Ong, Emily F Brownson, Iona AM Campbell, Craig Mowat, John P Seenan, Jonathan C MacDonald, Gwo-Tzer Ho

## Abstract

Extreme fatigue is a clinical symptom that affects >50% of individuals with Inflammatory Bowel Disease (IBD), with a similar prevalence across many common immune-mediated inflammatory diseases (IMIDs). Despite its ubiquity, human scientific studies have yet to explain the mechanistic basis of this pervasive and complex symptom. One fundamental reason for this, is our inability to account for the clinical heterogeneity of fatigue with its multifactorial nature. We present the conceptual machine learning (ML) framework to dissect the complex nature of fatigue using one of the largest prospectively captured, real-world patient-reported outcome (PROs) on wellbeing from three contemporaneous cohorts (2020-present), totalling 2,970 responses from 2,290 participants across the UK and internationally, including non-IBD controls with 100 lines of clinical metadata. We systematically defined the (1) threshold of fatigue as our primary outcome (≥10/14 fatigue days) to build our ML approach, (2) utilised routinely available clinical data that can be used at a population-level analysis, (3) employed seven different ML methods with external validation in 3 different cohorts in UK, Spain and Australia (n=252), (4) employed Shapley Additive Explanations (SHAP) analysis to break down the clinical heterogeneity to allow the examination of clinical predictive factors at an individual level; and finally (5), investigate whether there are distinct clusters of fatigue patients. We found that ML models performed comparably (AUC/C-index ∼0.7) on external validation with SHAP analysis showing interpretable, individualised fatigue drivers and five distinct fatigue phenotypes, including a subgroup of young males with significantly lower fatigue burden. Our data therefore provides the ML ‘roadmap’ to predict and deconstruct fatigue in IBD and potentially also more widely in IMIDs, enabling patient-level dissection beyond symptom-based classification with the ability to integrate deep molecular data. This is a step towards future clinical-scientific AI models with the immediate clinical application to stratify patients to human experimental studies to better understand the dominant mechanisms that drive fatigue at an individual level.

## Introduction

Many patients with Inflammatory Bowel Diseases (IBD) suffer from extreme fatigue, even when in remission^1–3^. From a patient’s perspective, fatigue is a major priority area for further research^4^. Extreme fatigue is also a common symptom across many immune-mediated inflammatory conditions (IMIDs) such as rheumatoid arthritis (RA), systemic lupus erythematosus (SLE) and sarcoidosis^5–7^. This suggest there may be unrecognised cross disease-mechanisms that are potentially independent of organ-specific inflammation^8^ and hence also poorly treated by conventional immune-suppression. There are increasingly powerful scientific tools from functional neuroimaging to metabolic assays to study the mechanistic basis of this pervasive symptom in IMIDs^9,10^. However, human experimental and interventional studies have been stymied by patient heterogeneity. Pertinently in IBD, few interventional studies have specifically targeted fatigue, and those that have, often show poor results^11–13^, are small and open-labelled in design^14^, and include ill-defined patient groups with fatigue. Our aim is to develop a machine-learning (ML) approach that can dissect the clinical heterogeneity of this complex symptom construct of fatigue. Here, we present an end-to-end machine learning roadmap in IBD utilising routinely available clinical and laboratory data to do this at a population level whilst retaining the ability to investigate characteristics on the individual level with the objective of deeper patient stratification - ‘to find the right patients, for the right studies’.

## Methods

### Patient data

We utilised patient-reported outcome (PRO) fatigue data from two prospective mechanistic biomarker studies, namely (1) Investigation into Gastrointestinal Damage Associated Molecular Patterns (GI-DAMPs) that is a cross-sectional IBD study with ethical approvals from the East Scotland Ethics Committee (REC 18/ES/0090); and (2) Mitochondrial DAMPs as mechanistic biomarkers of mucosal inflammation in Crohn’s Disease (MUSIC study; www.musicstudy.uk), a prospective IBD cohort study with ethical approvals by East Scotland Ethics Committee (REC 19/ES/0087). Both are cohort studies carried out in Scotland (Glasgow, Edinburgh and Dundee; 2020-present) that recruit participants with IBD along with ∼100 lines of clinical metadata including IBD activity, treatment, comorbidities and laboratory parameters. In addition to this, we conducted an online survey to further collect PRO data on fatigue (2023-2024; n=1,643 within UK, n=112 internationally).

In all 3 studies (Table 1), we used the validated Crohn’s and Ulcerative Colitis Questionnaire-32 (CUCQ32) questionnaire that is applicable to Crohn’s disease (CD) and Ulcerative colitis (CD), the 2 sub-types of IBD^15^. CUCQ32 contains 32 questions that measures 4 domains of wellbeing (gastrointestinal, social, psychological and general wellbeing) with 5 main questions pertaining to fatigue, generating a score ranging from 0-272. Collectively, these studies involved 2,290 total participants (including 336 non-IBD participants who responded online, and 27 non-IBD symptomatic controls) and provide the scale of data to establish a clinical threshold of patient-reported fatigue as a baseline for our ML approach.

**Table 1:**
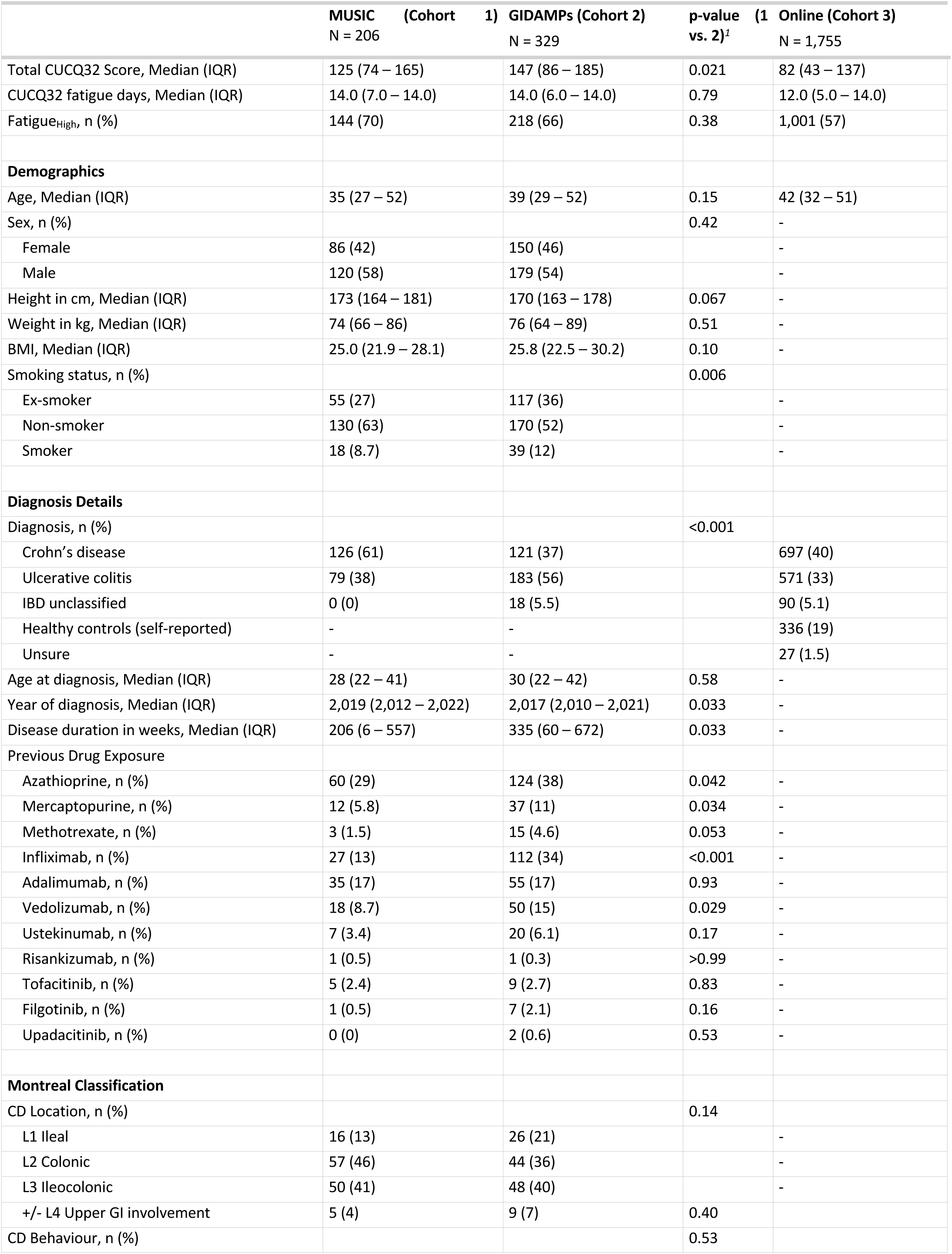
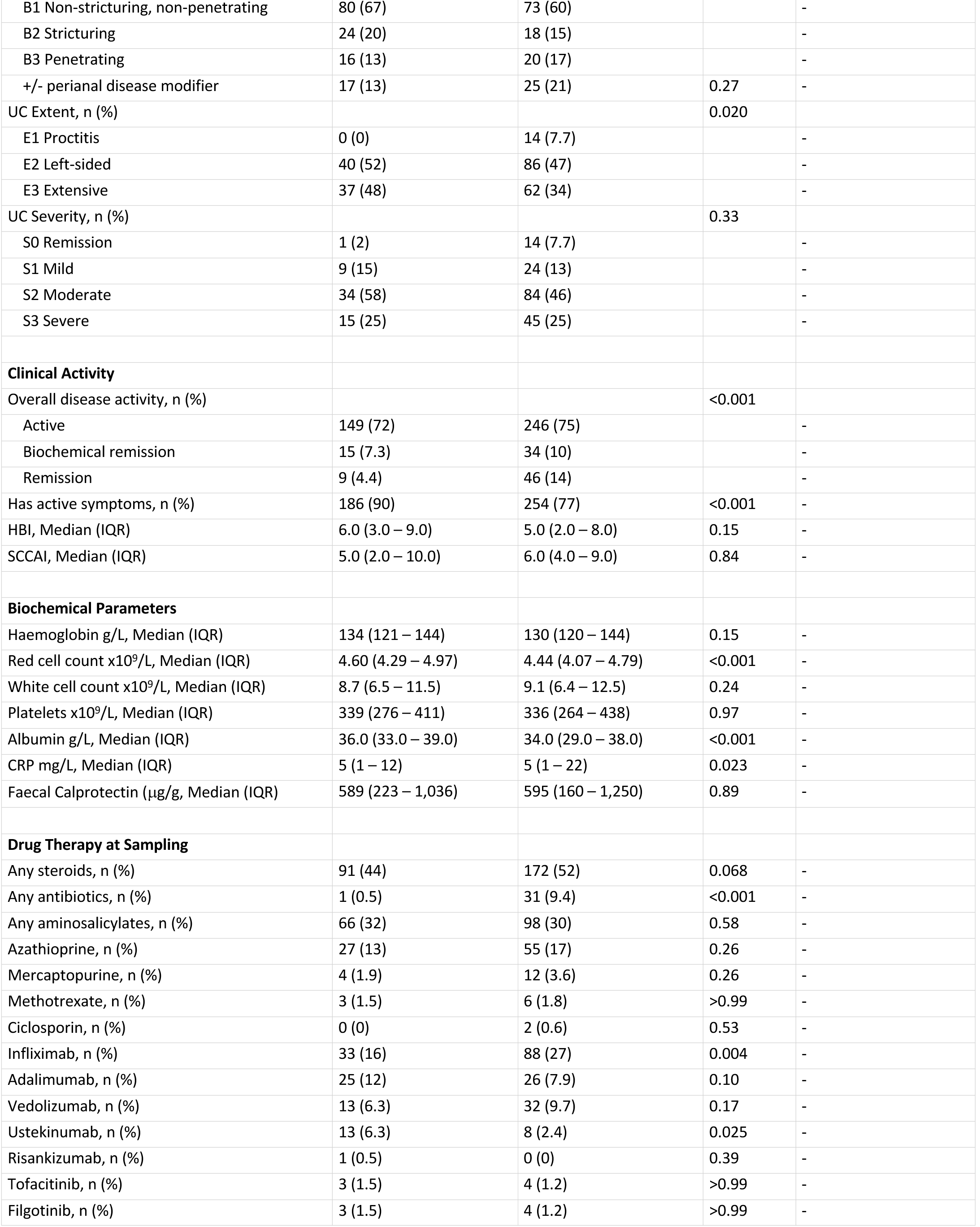
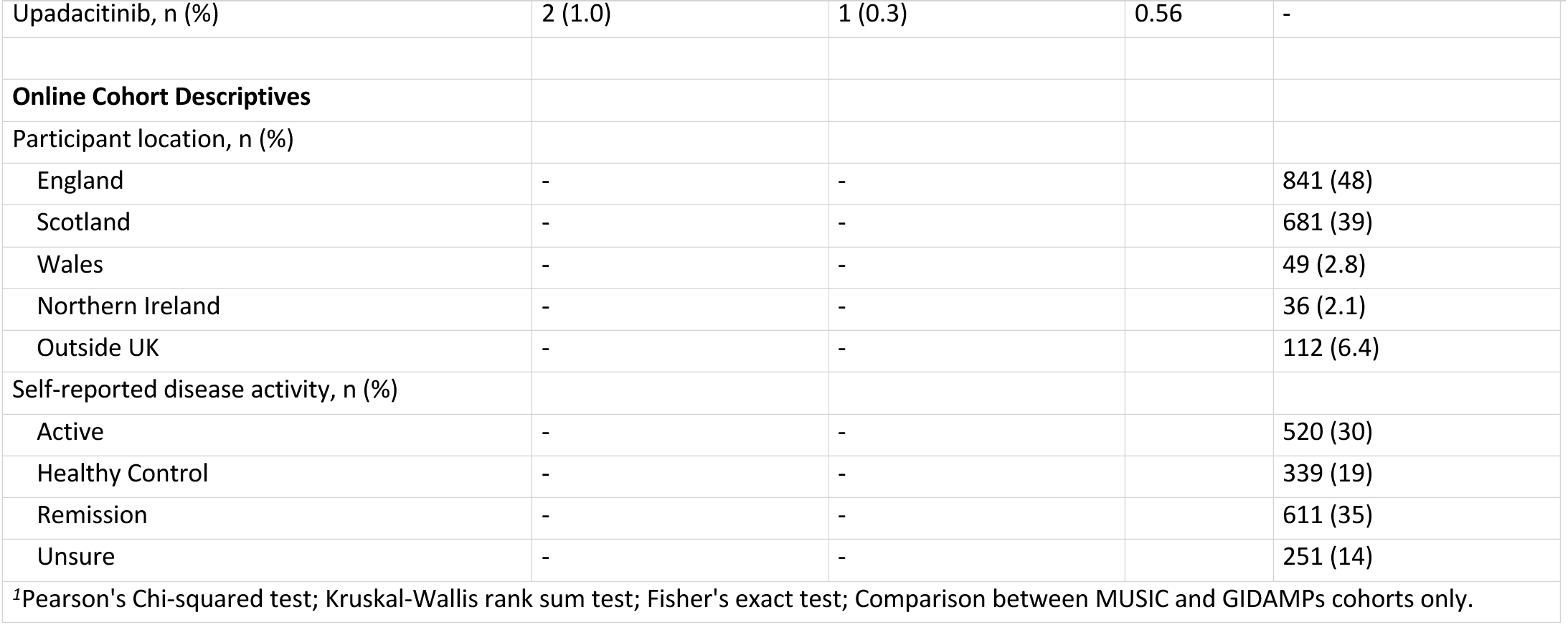
Demographics and clinical characteristics of Cohorts 1 (MUSIC), 2 (GI-DAMPs) and 3 (self-reported online).

### Model development and evaluation of machine learning pipeline

Input features include clinical (e.g. diagnosis group, Montreal classification), demographics, exposome (smoking, alcohol, seasonality), laboratory (e.g. CRP, faecal calprotectin) and IBD drug exposure data (Full list in Supplementary Table 3). Seven supervised ML models were employed: XGBoost, Random Forest, AdaBoost, multilayer perceptron (MLP), support vector machine, logistic regression (*scikit-learn* implementation), and a custom feedforward deep neural network implemented in TensorFlow (Supplementary Figure 4c). Conventional logistic regression (*statsmodels*) was included as a reference comparator. Analyses were conducted using Python (version 3.11.9), with the following libraries: *scikit-learn* (v1.5.2), *scipy* (v1.14.1), *xgboost* (v2.1.2), *pandas* (v2.2.3), *numpy* (v2.0.2), *shap* (v0.46.0), *tensorflow* (v2.18.0), *pytorch* (v2.5.1), *statsmodels* (v0.14.4), *seaborn* (v0.13.2), and *matplotlib* (v3.10.0).

Missing data were imputed according to variable type. Continuous variables with non-normal distributions (e.g., C-reactive protein (CRP), albumin, faecal calprotectin) were imputed using the median to minimise bias. Categorical variables were transformed using one-hot encoding. Numerical variables were standardised (zero mean, unit variance) using *StandardScaler* (*scikit-learn*). Derived features included seasonality (calculated from the date of CUCQ32 completion) and disease duration (expressed in weeks since diagnosis). To avoid information leakage from repeated measures, group-aware data splitting was performed using *GroupKFold* and *GroupShuffleSplit* (*scikit-learn*), with participant identifiers used as grouping variables. For conventional ML models, data were partitioned into training-validation (80%) and independent testing (20%) sets, with hyperparameter tuning conducted via 5-fold group cross-validation. For deep neural networks, data were split into training (65%), validation (15%), and testing (20%) sets using the same group-aware strategy.

Model interpretability was evaluated using Shapley Additive Explanations (SHAP)^16^. Model generalisability was evaluated using fully anonymised datasets from three prospectively collected independent validation cohorts based in Scotland, Spain, and Australia (Supplementary Table 5, predominantly recruited in the outpatient IBD clinic setting). These datasets were entirely independent of model training and hyperparameter optimisation. Identical preprocessing steps were applied to the external datasets prior to prediction. Model performance was assessed using receiver operator (ROC) curves, area under the curve (AUC), sensitivity, specificity, positive and negative predictive values (PPV and NPV respectively).

Unsupervised clustering was performed using K-means. Following exclusion of a single outlier datapoint, the optimal number of clusters was empirically determined by evaluating solutions for k = 2 to k = 10, informed by prior clinical knowledge that the cohort contained a subgroup with active IBD. A solution with k = 5 was selected, balancing cohort heterogeneity while avoiding over-fragmentation or generation of spurious small clusters.

All anonymised datasets, dependencies, and code to reproduce this study are publicly available at: https://github.com/1-gut/machine_learning_for_ibd_fatigue.

## Results

### Defining the threshold of fatigue

We modelled our ML algorithms to predict IBD patients with patient-reported outcome fatigue of ≥10 days over the past 14 days at time of data entry, as the primary outcome variable (Fatigue PRO). This is based on CUCQ32 question: “On how many days over the last 2 weeks did you feel tired?” (0–14 days). Across the combined dataset, IBD patients with active disease reported significantly more fatigued days than those in remission (medians 14 vs 7 days; p<0.001). In IBD patients in remission, fatigue days remained significantly elevated compared to non-IBD controls. (7 vs 4 days; p<0.001; Supplementary Figure 1a). These findings were consistent across individual cohorts. Fatigue days were strongly correlated with total CUCQ32 scores (r=0.73, p<0.0001; Supplementary Figure 1b). The distribution of fatigue days was skewed (Supplementary Figure 1c). Therefore, we used the median value of ≥10 days of fatigue over 14 days and defined it as the threshold for clinically significant fatigue (Fatigue_high_). Using this definition, 53.9% (1602/2970) of observations met criteria for Fatigue_high_, comprising 75.2% of active IBD patients, 43.6% of those in remission, and 14.2% of non-IBD controls. Patients in the Fatigue_high_ group had significantly higher overall CUCQ32 scores than those below this threshold (median 130 vs 38; p<0.0001; Supplementary Figure 1d).

Fatigue PRO is highly correlated to all measures of CUCQ32 domains (all p<0.001). This shows that a simple question is relevant in the complex construct of fatigue. PROs (‘*what patients tells clinicians*’) are strongly correlated with clinician-based assessment (‘*what clinicians think of patients*’) and are associated with clinical parameters such as CRP and faecal calprotectin (Supplementary Figure 2, both p<0.0001). Among 147 MUSIC patients with longitudinal follow-up over 12 months, serial CUCQ32 measurements demonstrated heterogeneity between individuals. Patients achieving clinical remission (Harvey Bradshaw Index, HBI≤5 for CD and Simple Clinical Colitis Activity Index SCCAI≤2 for UC) at 12 months tended to show improvements in both overall CUCQ32 scores and fatigue-specific domains (Supplementary Figure 3). However, no significant difference in CUCQ32 was observed between those with or without mucosal healing at 12 months (median 68 vs 62; Supplementary Figure 2e), indicating that endoscopic remission does not fully capture patient-reported wellbeing. Collectively, this demonstrates the clinical heterogeneity of fatigue when assessed in accordance with conventional measures of inflammation in IBD.

### Development of machine learning algorithms and external validation to predict fatigue

Having defined our threshold for fatigue, we incorporated self-reported fatigue assessments and all available clinical parameters such as disease activity indices (HBI and SCCAI), laboratory parameters (C-Reactive Protein [CRP], faecal calprotectin, full blood count), body mass index (BMI), smoking status, drug therapy, and seasonality. Here, we generated an initial training dataset of 1,215 observations from Cohorts 1 and 2 for seven ML algorithms: XGBoost, random forest, AdaBoost, multilayer perceptron classifier, support vector classifier, logistic regression, and a custom-built deep neural network (DNN). *GroupKFold* cross-validation ensured that repeated measures from individual patients remained in the same dataset split. Across classical ML models, predictive performance was broadly comparable with AUC values ranging from 0.69-0.73 for Fatigue_high_ prediction (Figure 1b). To explore whether complex nonlinear patterns might enhance predictive accuracy, we developed a custom Deep Neural Network (DNN) implemented in TensorFlow and re-coded in PyTorch. The network architecture comprised four layers (two dense layers interspersed with two dropout layers; Supplementary Figure 4c). On internal test data, the DNN outperformed classical ML models, achieving an AUC of 0.89 for all IBD cases and 0.94 in patients in biochemical remission (Figure 1b). However, external validation using an independent cohort (n = 252; Córdoba, Spain n = 101; Melbourne, Australia n = 90; Edinburgh, Scotland n = 61) revealed that the DNN’s performance did not generalise, with AUCs falling back to the 0.69-0.73 range observed across all models (Figure 1c). This suggests that the superior internal performance likely reflected model overfitting rather than true generalisability. No single ML model demonstrated consistent superiority across all datasets.

**Figure 1.**
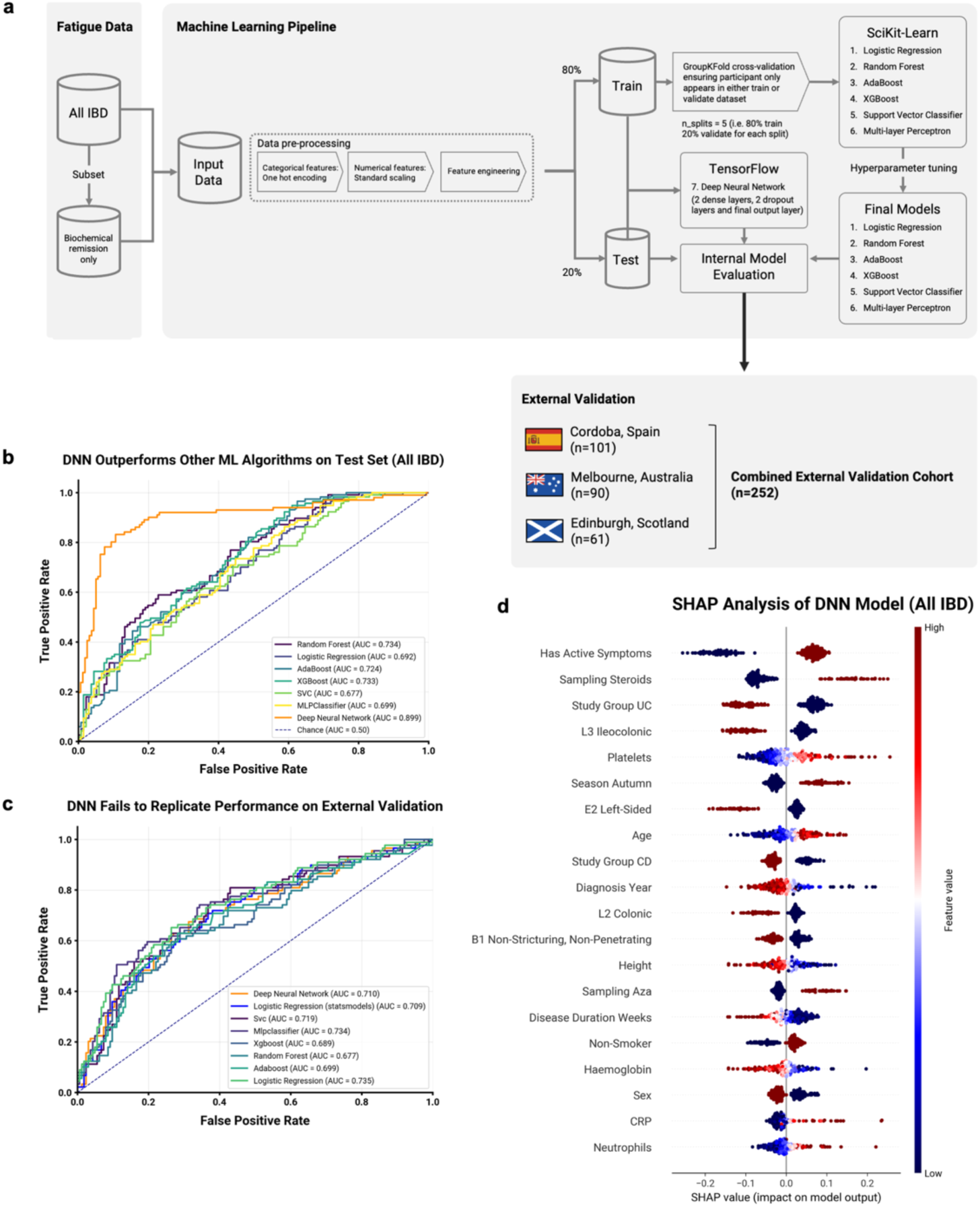
Machine learning workflow, performance, and interpretability of models predicting fatigue in IBD patients. (a) Overview of the machine learning workflow. Two pipelines were implemented: one including all IBD patients (Cohorts 1-2), and another restricted to patients in biochemical remission (defined as calprotectin <250 µg/g and CRP <5 mg/dL). Model performance was subsequently evaluated on an external validation cohort comprising patients with IBD from Spain, Australia, and Scotland. (b) Model performance (AUG) in the full IBD cohort. The deep neural network (DNN) achieved the highest test set performance compared to other models, motivating external validation. (c) Model performance (AUC) in the external validation cohort. All models performed similarly, though the DNN showed a notable decrease in AUC, suggesting possible overfitting in the initial test dataset. (d) SHAP summary plot for the DNN, illustrating the impact of individual features on model predictions. Each dot represents a patient’s data point for a specific feature. The horizontal position of the dot shows how much that feature pushes the prediction toward fatigue (positive values) or away from fatigue (negative values) Dot color reflects the actual feature value (e.g., red for high platelet counts, blue for low). Features are listed vertically in order of their overall impact on the model, with the most influential features at the top. This visualization reveals which features most strongly influence fatigue predictions and highlights that the impact of features can vary between patients—for example, very high platelet counts increase predicted fatigue in some individuals but not in others. Assessments conducted in autumn also increased the likelihood of fatigue prediction.

When restricting the analysis to patients in biochemical remission (CRP < 5 mg/L and calprotectin < 250 μg/g), model performance declined (AUCs 0.61-0.66 (Supplementary Figure 4a). In this subgroup, inflammatory markers contributed less to fatigue prediction, while features such as anaemia, elevated lymphocytes, reduced urea (potentially reflecting sarcopenia), age, and seasonality emerged as more influential (Supplementary Figure 4b).

### Personalised Fatigue Profiling Using SHAP-Driven Model Interpretation

To better understand how individual factors contributed to model predictions, we applied SHAP analysis^16^. SHAP values quantify the relative contribution of each input variable to individual model predictions, offering a transparent, patient-specific decomposition of the model’s decision-making process. SHAP interpretation revealed distinct patient-level patterns, underscoring the clinical (and possibly biological) heterogeneity of fatigue in IBD. For some individuals, fatigue was closely linked to inflammatory markers, while for others, factors such as weight, medication use, or seasonal variation were more influential. In the DNN model, key predictive features for all IBD patients included clinical symptom activity, steroid use, IBD diagnosis (CD vs UC), platelet count, and autumn season (Figure 1d). By enabling granular, patient-specific insights into the drivers of fatigue, SHAP analysis provides a proof-of-concept pathway toward personalised mechanistic understanding of fatigue in IBD (Figure 2).

**Figure 2.**
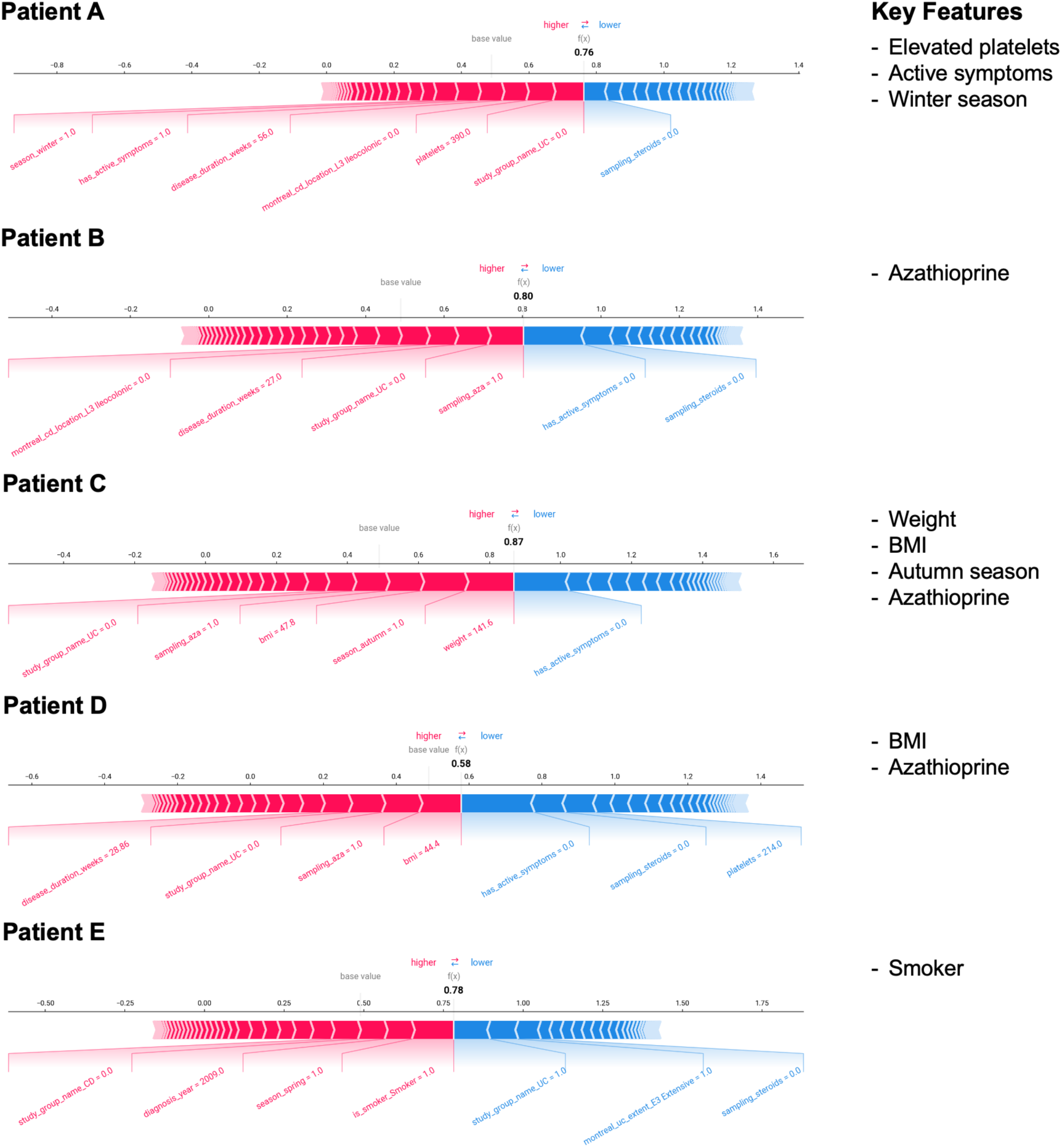
Individualised decomposition of IBD-related fatigue using SHAP. SHAP force plots for true positive cases (i.e., instances where the patient reports fatigue and the model predicts fatigue). f(x) represents the model-predicted probability of Fatigue_High_; for example, f(x) = 0.76 corresponds to a 76% probability of Fatigue_High_· Values closer to 1 indicate higher model confidence in predicting fatigue, while values closer to O indicate confidence in predicting low fatigue. This proof of concept illustrates the model’s ability to decompose feature contributions at the individual level. For example, in patient A, active IBD is a major contributor to fatigue; in patient B, azathioprine is a more influential factor. Patients C and D exhibit multifactorial contributions, with an important weight-related component, while patient E’s smoking status emerges as a key driver of fatigue.

### Identification of Distinct Fatigue Phenotypes Using K-Means Clustering

To further characterise potential fatigue subtypes, we applied unsupervised k-means clustering (Figure 3). Five distinct patient clusters emerged (Table 2) namely those with (1) Active IBD, (2) Young tall males, (3) Crohn’s disease patients with long disease duration, (4) More elderly UC patients; and (5) Young female IBD patients. Cluster 0, representing active IBD, exhibited the highest prevalence of Fatigue_high_ (81.4%, n=113). In contrast, Cluster 1 (young tall males) demonstrated a significantly lower fatigue prevalence (34.9%) compared to all other groups (ANOVA F=22.6, p<0.0001; Tukey HSD post hoc comparisons; Figure 4c). The remaining clusters exhibited intermediate fatigue prevalence ranging from 50–56%. The low-fatigue male cluster (Cluster 1) was distinct not only in sex (96% male), height (median 1.80 m), and weight (median 84 kg), but also exhibited higher haemoglobin levels (145 g/L vs 120-137 g/L in other clusters). The reasons for their relative fatigue resistance remain unclear but may reflect biological resilience, under-reporting, or unidentified protective mechanisms. Notably, deviations from low baseline fatigue within this group may represent meaningful signals warranting focused mechanistic investigation.

**Figure 3.**
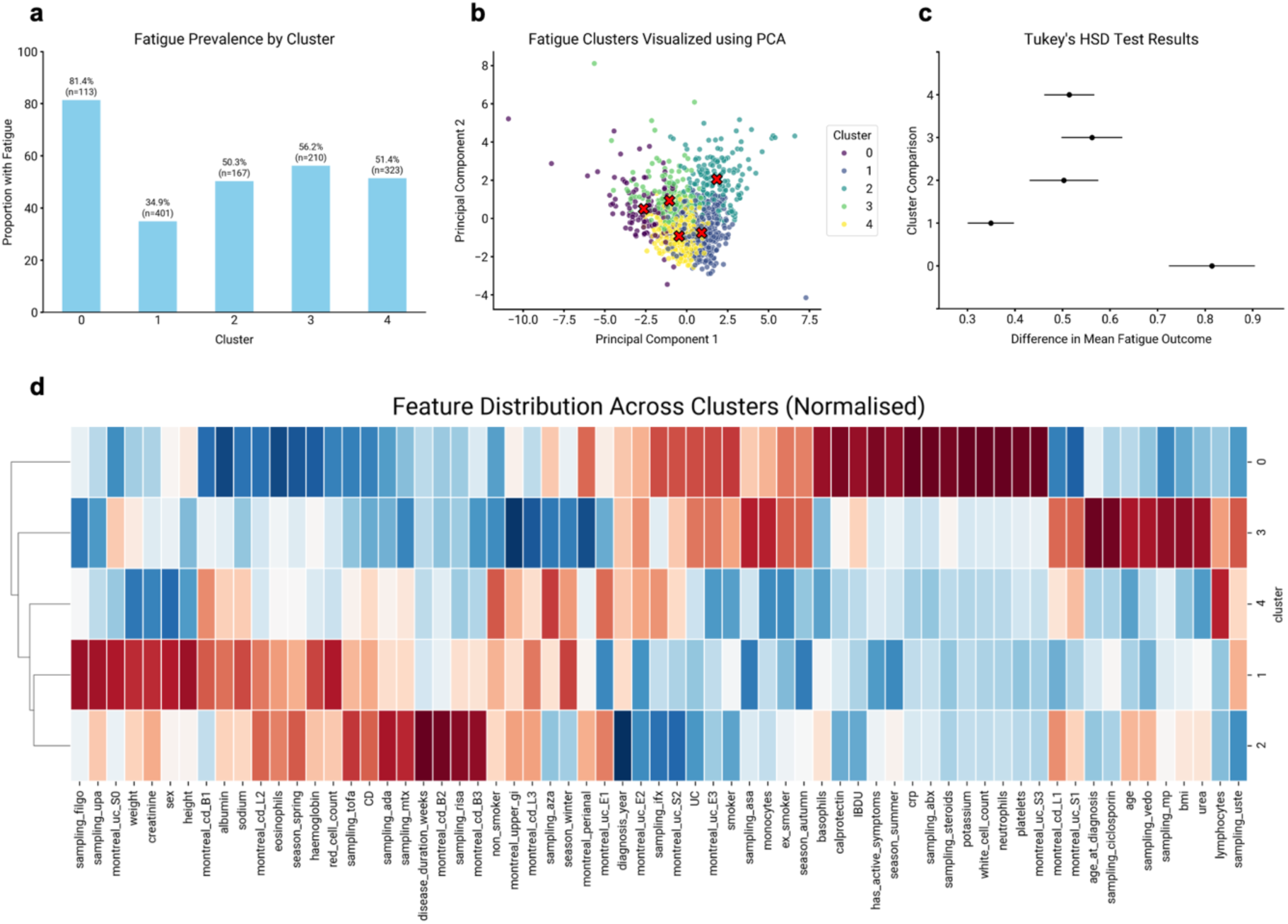
K-means clustering identifies five distinct fatigue clusters. Fatigue prevalence by cluster: cluster O shows high fatigue prevalence (81%), cluster 1 shows low fatigue prevalence (35%), and clusters *2--4* show intermediate fatigue prevalence (50-56%). (a) Visualization of fatigue clusters using principal components analysis (PCA). (b) Tukey’s HSD test indicates that cluster O has significantly higher fatigue compared to the other clusters, while cluster 1 has significantly lower fatigue. No significant differences are observed among clusters *2--4*. (c) Heatmap visualization of the five identified subgroups.

**Table 2.**
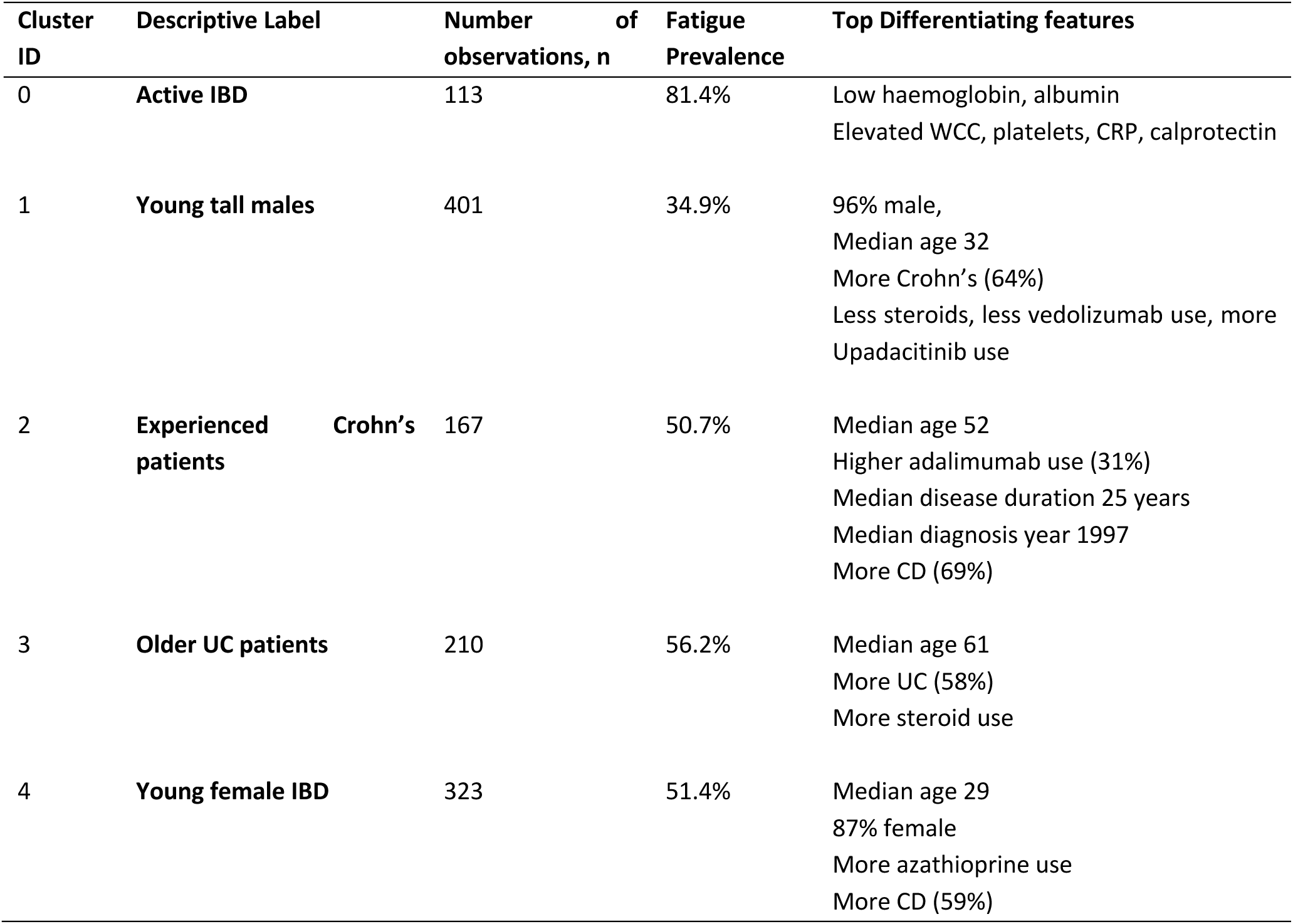
Summary Cluster Characteristics and Descriptive Label.

Together, these findings demonstrate that IBD-associated fatigue remains prevalent despite disease control, exhibits marked individual variability and can be partially modelled using supervised and unsupervised ML approaches that integrate multidimensional clinical data.

## Discussion

Artificial intelligence and powerful machine learning are now firmly established and are increasingly utilised in medicine^17^. A key consideration is how we construct the practical steps and apply these tools to a complex and relevant medical patient-centric problem, such as fatigue. Here, we present the conceptual ML framework ‘roadmap’ to dissect the fatigue towards a practical outcome of better patient stratification to aid mechanistic studies; using one of the largest prospectively captured, real-world PROs on wellbeing from three contemporaneous cohorts, totalling 2,970 responses from 2,290 participants across the UK and internationally, including non-IBD controls. We systematically defined the relevance of the (1) primary outcome of fatigue to based our ML approach, (2) utilised routinely available clinical data that can be used at a population-level analysis, (3) employed 7 different ML methods, (4) employed SHAP analysis to break down the clinical heterogeneity at an individual level; and finally (5), investigate whether there are distinct clusters of fatigue patients based simply on routine clinical observations.

We found that our ML approach can predict IBD patients with fatigue (AUC 0.70) with external validation across 3 independent cohorts in different geographical locations, but the performance drops within the IBD patient group that is in remission. We systematically compared seven ML models against traditional logistic regression (Appendix 4) to evaluate their ability to incorporate deep clinical data. Deep neural networks demonstrated superior internal performance but similar external validation performance across all models. However, substantial unexplained variance remains, particularly in remission.

The use of our simplified outcome threshold of ≥10/14 fatigue days deserves further discussion. Firstly, the prevalences of fatigue defined by this agrees with clinical studies of IBD and IMID fatigue^18,19^. Secondly, this allows us to harmonise fatigue measurement across cohorts, scale up our dataset and facilitates predictive machine learning analyses that can be applicable to different IMID cohorts in the future. Thirdly, our data from the broader CUCQ32 shows that this simple PRO is highly correlated to other domains of wellbeing and therefore, a good representation of the deeper construct of the symptom fatigue that may be linked to poor sleep, depression, anxiety and organic disease-related causes. Hence, our goal is to build a ML-approach that can capture these complexities from a simple starting point that is easily captured in the clinic.

A potential critique is that fatigue prediction models may merely recapitulate the fatigue question itself i.e. reinforcing a circular argument. We address this by explicitly excluding the fatigue question and other CUCQ questions as input variables in our ML algorithms. Instead, fatigue was predicted from a broad range of independently collected clinical metadata, including laboratory markers (e.g., CRP, calprotectin), medication exposure, BMI, smoking status, and seasonality. These predictors are biologically and clinically orthogonal to self-reported fatigue. Furthermore, SHAP analysis revealed interpretable, participant-specific predictors such as inflammatory markers and smoking, providing face validity to the modelling. External validation across independent cohorts confirmed that fatigue can be predicted to an extent without direct fatigue self-report, demonstrating the capacity of machine learning to uncover latent patterns underlying fatigue.

This suggests that (1) machine learning is equivalent to classical methods for structured clinical data; and (2) the consistent performance ceiling likely reflects a ‘hidden’ multifactorial pathobiological compartment of factors currently unmeasured. We, therefore, present the first comprehensive framework utilising ML-algorithms to dissect the complex presentation of fatigue. This is initially based on using routinely available clinical data but molecular data such as genetics, nutritional factors, microbiome and metabolomics for examples, can be easily incorporated. We acknowledge that our dataset lacked key variables known to influence fatigue, including sleep quality (we excluded the use of CUCQ sleep question as it was highly correlated to the outcome), mental health, socioeconomic status, and more detailed comorbidities. We think that with further training of the models, there will be direct clinical application by identifying ‘clusters of patients’ or individualised stratification based on SHAP analysis to human experimental studies of fatigue. These insights may ultimately support patient stratification to targeted interventional studies.

In conclusion, fatigue profoundly impacts wellbeing in IBD, even in remission. Our data provides credible support for the utility of patient reported outcomes as endpoints for future translational scientific research. In addressing IBD-associated fatigue, we show the conceptual ability to reduce the dimensionality via modelling (thus flattening out the heterogeneity of IBD-associated fatigue) as the first-pass mechanism to identify fatigue subgroups. This simplified approach will allow the future incorporation of multiple streams of complex scientific metadata (such as genetics and microbiome for example) for larger scale analyses at a cohort level. There are many better scientific tools to study central and peripheral fatigue - from imaging, metabolism and mitochondrial function. We envisage that our work provides a step towards identifying clear subgroups for targeted mechanistic studies and therapeutic development, shifting beyond symptom-based classification towards data-driven and personalised fatigue management in IBD.

## Data Availability

All data produced are available online at https://github.com/1-gut/machine_learning_for_ibd_fatigue

## Acknowledgements

This work is part of the MUSIC IBD study, funded by The Leona M. and Harry B. Helmsley Charitable Trust (G-1911-03343) and the Anne Ferguson Memorial Fund to GTH.

## Supplementary Material

**Supplementary Table 1:** The CUCQ32 questions.

|  | Questions |
| --- | --- |
| 1 | On how many days over the last two weeks have you had loose or runny bowel movements? |
| 2 | On how many days over the last two weeks have you felt generally unwell? |
| 3 | On how many days over the last two weeks have you had to rush to the toilet? |
| 4 | On how many nights in the last two weeks have you had to get up to use the toilet because of your bowel condition after you have gone to bed? |
| 5 | On how many days over the last two weeks have you felt tired? |
| 6 | On how many days over the last two weeks have you wanted to go back to the toilet immediately after you thought you had emptied your bowels? |
| 7 | In the last two weeks have you felt the need to keep close to a toilet? |
| 8 | On how many days over the last two weeks have you felt pain in your abdomen? |
| 9 | On how many days over the last two weeks has your abdomen felt bloated? |
| 10 | In the last two weeks did your bowel condition prevent you from going out socially? |
| 11 | On how many days over the last two weeks have you opened your bowels more than three times a day? |
| 12 | On how many nights over the last two weeks have you been unable to sleep well (days if you are a shift worker)? |
| 13 | In the last two weeks, has your bowel condition prevented you from carrying out your work or other normal activities? |
| 14 | In the last two weeks, has your bowel condition affected your leisure or sports activities? |
| 15 | In the last two weeks have you felt irritable? |
| 16 | On how many days over the last two weeks have you felt full of energy? |
| 17 | On how many days in the last two weeks have you noticed blood in your stools? |
| 18 | On how many days over the last two weeks have you felt off your food? |
| 19 | In the last two weeks, has your sex life been affected by your bowel problem? |
| 20 | On how many days over the last two weeks, have you had a problem with large amounts of wind? |
| 21 | In the last two weeks have you felt angry as a result of your bowel problem? |
| 22 | On how many days over the last two weeks have your bowels opened accidentally? |
| 23 | In the last two weeks have you felt relaxed? |
| 24 | In the last two weeks have you felt depressed? |
| 25 | On how many days over the last two weeks have you felt sick? |
| 26 | In the last two weeks have you had to avoid attending events where there was no toilet close at hand? |
| 27 | In the last two weeks have you felt frustrated? |
| 28 | In the last two weeks have you been embarrassed by your bowel problem? |
| 29 | Many patients with bowel problems have worries about their illness. How often during the last two weeks have you felt worried? |
| 30 | In the last two weeks have you felt upset? |
| 31 | In the last two weeks have you felt happy? |
| 32 | In the last two weeks have you felt lack of sympathy from others? |

**Supplementary Table 2:** Paired correlation analyses of CUCQ32 questions against question 5, (‘How many days in the last 2 weeks did you feel tired?’). Spearman-rank analyses, Benjamini-Hochberg corrected.

| CUCQ32 Question Number | How many days are you tired in the last 2 weeks (Q5)? |
| --- | --- |
| 1 | 1.33E-69 |
| 2 | 3.19E-147 |
| 3 | 2.01E-85 |
| 4 | 1.40E-51 |
| 6 | 3.90E-67 |
| 7 | 1.96E-82 |
| 8 | 2.44E-102 |
| 9 | 7.58E-93 |
| 10 | 3.85E-86 |
| 11 | 1.16E-65 |
| 12 | 4.80E-103 |
| 13 | 6.59E-79 |
| 14 | 3.45E-90 |
| 15 | 2.03E-82 |
| 16 | 3.25E-142 |
| 17 | 1.89E-35 |
| 18 | 8.76E-67 |
| 19 | 1.23E-63 |
| 20 | 1.39E-46 |
| 21 | 7.60E-47 |
| 22 | 3.08E-21 |
| 23 | 3.07E-49 |
| 24 | 1.91E-48 |
| 25 | 5.80E-57 |
| 26 | 1.47E-47 |
| 27 | 4.80E-73 |
| 28 | 1.26E-53 |
| 29 | 1.73E-57 |
| 30 | 8.37E-63 |
| 31 | 1.61E-28 |
| 32 | 2.97E-09 |

**Supplementary Table 3:**
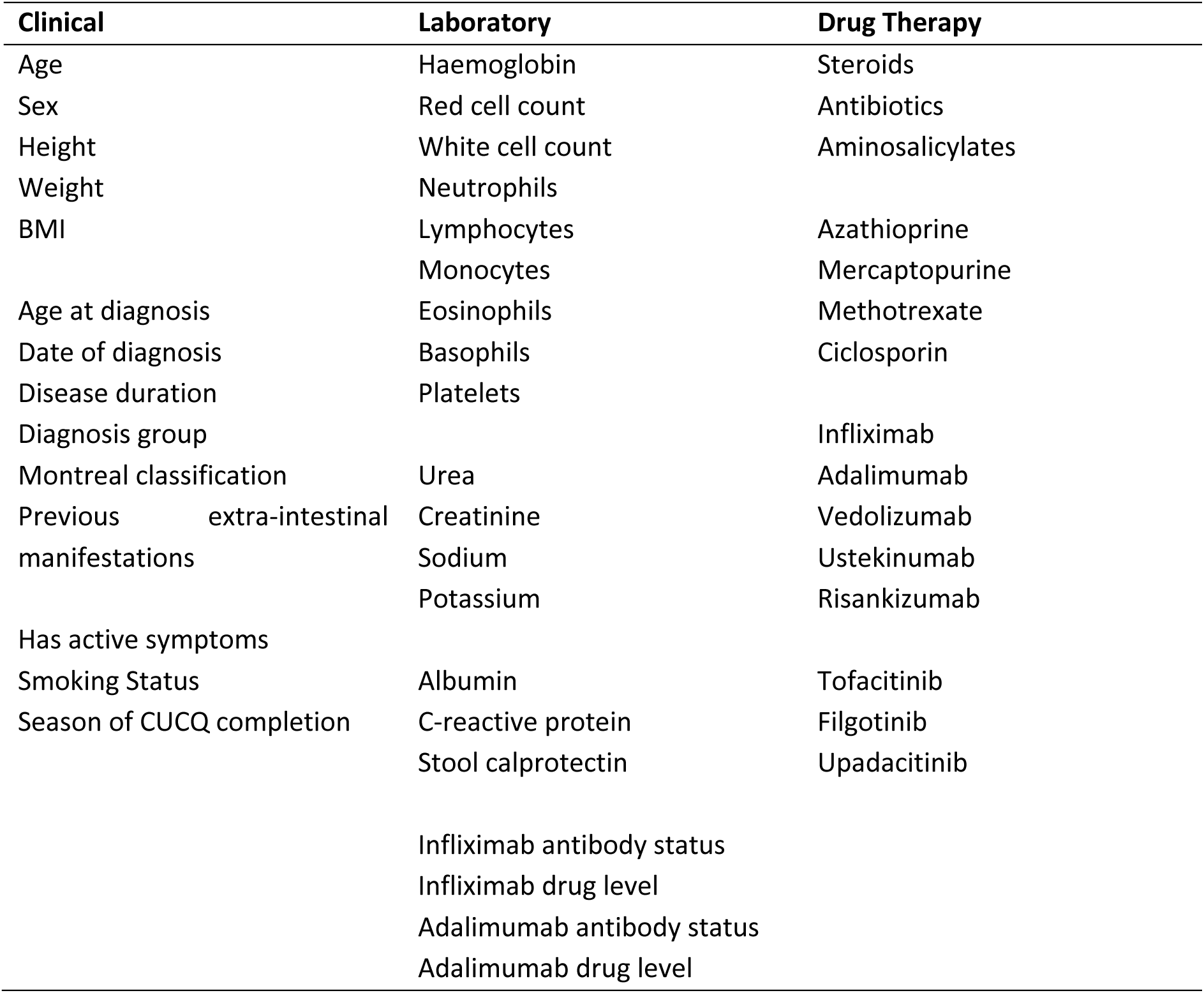
List of input features for machine learning after data transformation.

**Supplementary Table 4:**
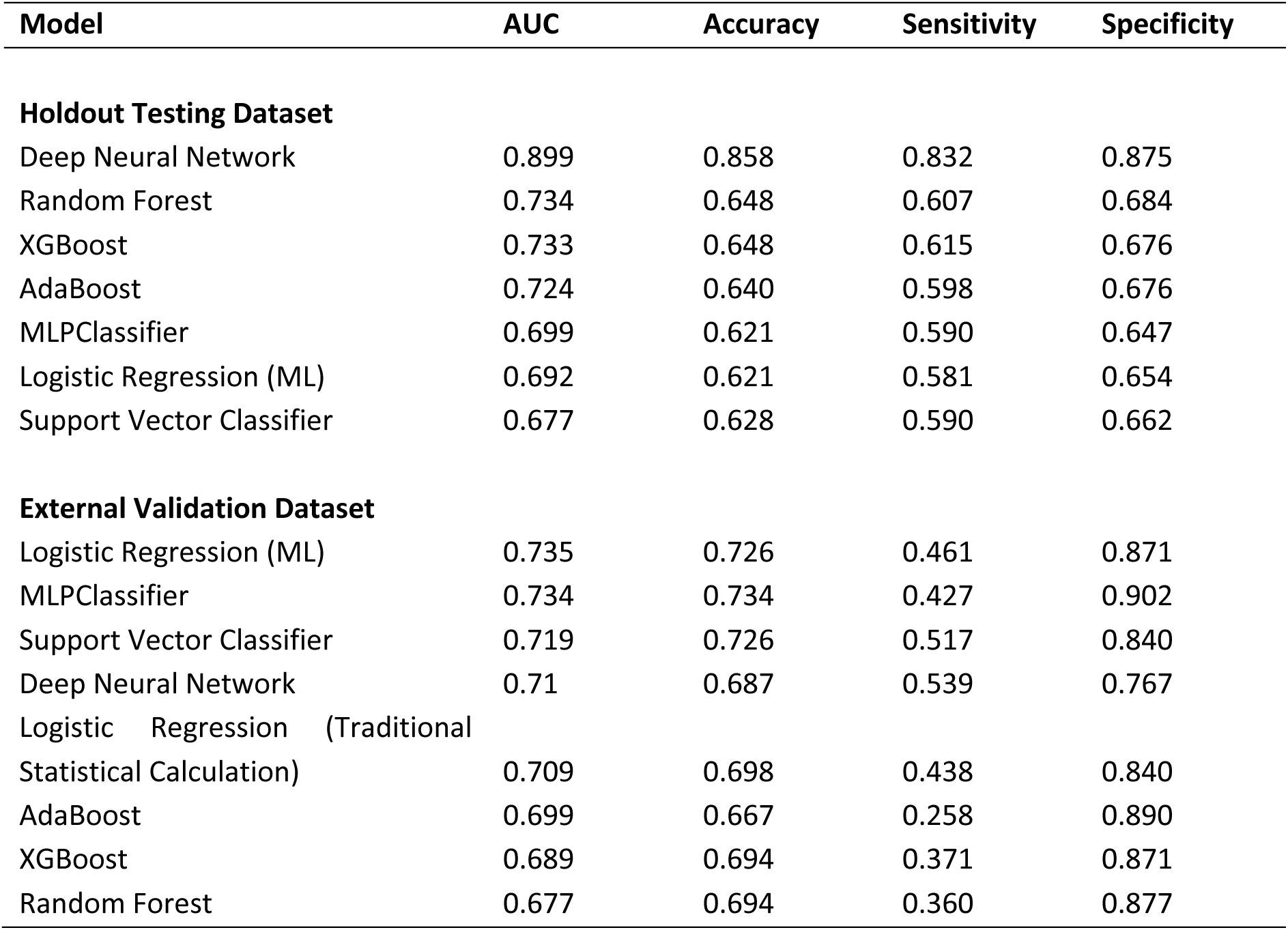
Model metrics for the 7 different machine learning approaches (All IBD) ranked by area under receiver-operator curve (AUC)

**Supplementary Table 5:** Summary of Validation Cohorts.

| Characteristic | Overall N = 252 | Australia N = 90 | Scotland N = 61 | Spain N = 101 | p-value<sup>†</sup> |
| --- | --- | --- | --- | --- | --- |
| Fatigue<sub>High</sub>, n (%) | 89 (35) | 35 (39) | 33 (54) | 21 (21) | <0.001 |
| **Demographics** |  |  |  |  |  |
| Age, Median (IQR) | 41 (31 – 57) | 37 (26 – 51) | 43 (34 – 62) | 42 (31 – 57) | 0.030 |
| Sex, n (%) |  |  |  |  | 0.015 |
| Female | 122 (48) | 42 (47) | 39 (64) | 41 (41) |  |
| Male | 130 (52) | 48 (53) | 22 (36) | 60 (59) |  |
| Height in cm, Median (IQR) | 170 (163 – 175) | 172 (164 – 172) | 169 (160 – 176) | 169 (163 – 175) | 0.65 |
| Weight in kg, Median (IQR) | 76 (65 – 88) | 75 (64 – 90) | 82 (73 – 92) | 74 (64 – 84) | 0.018 |
| BMI, Median (IQR) | 25.5 (23.6 – 28.7) | 25.5 (24.5 – 26.4) | 27.3 (24.9 – 32.2) | 24.9 (22.4 – 27.5) | 0.002 |
| Smoking status, n (%) |  |  |  |  | <0.001 |
| Non-smoker | 146 (61) | 69 (77) | 37 (74) | 40 (40) |  |
| Ex-smoker | 61 (25) | 10 (11) | 9 (18) | 42 (42) |  |
| Smoker | 34 (14) | 11 (12) | 4 (8.0) | 19 (19) |  |
| **Diagnosis Details** |  |  |  |  |  |
| Diagnosis, n (%) |  |  |  |  | 0.41 |
| Crohn's disease | 158 (63) | 52 (58) | 39 (65) | 67 (66) |  |
| Ulcerative colitis | 87 (35) | 37 (41) | 20 (33) | 30 (30) |  |
| IBD unclassified | 6 (2.4) | 1 (1.1) | 1 (1.7) | 4 (4.0) |  |
| Age at diagnosis, Median (IQR) | 29 (21 – 42) | 27 (19 – 35) | 28 (22 – 43) | 32 (22 – 47) | 0.036 |
| Year of diagnosis, Median (IQR) | 2017 (2010 – 2023) | 2018 (2009 – 2023) | 2014 (2006 – 2018) | 2017 (2012 – 2023) | 0.011 |
| Disease duration in weeks, Median (IQR) | 445 (111 – 809) | 375 (117 – 849) | 584 (373 – 999) | 426 (66 – 690) | 0.008 |
| **Montreal Classification** |  |  |  |  |  |
| CD Location, n (%) |  |  |  |  | 0.19 |
| L1 Ileal | 57 (36) | 18 (34) | 9 (23) | 30 (45) |  |
| L2 Colonic | 34 (21) | 13 (25) | 10 (25) | 11 (17) |  |
| L3 Ileocolonic | 68 (43) | 22 (42) | 21 (53) | 25 (38) |  |
| +/- L4 upper GI involvement | 8 (3.2) | 0 (0) | 4 (6.6) | 4 (4.0) | 0.036 |
| CD Behaviour, n (%) |  |  |  |  | 0.49 |
| B1 Non-stricturing, non-penetrating | 104 (65) | 30 (57) | 30 (75) | 44 (66) |  |
| B2 Stricturing | 27 (17) | 11 (21) | 5 (13) | 11 (16) |  |
| B3 Penetrating | 29 (18) | 12 (23) | 5 (13) | 12 (18) |  |
| +/- perianal disease modifier | 25 (9.9) | 6 (6.7) | 7 (11) | 12 (12) | 0.43 |
| UC Extent, n (%) |  |  |  |  | 0.10 |
| E1 Proctitis | 17 (19) | 3 (7.9) | 6 (32) | 8 (25) |  |
| E2 Left-sided | 19 (21) | 11 (29) | 4 (21) | 4 (13) |  |
| E3 Extensive | 53 (60) | 24 (63) | 9 (47) | 20 (63) |  |
| UC Severity, n (%) |  |  |  |  | 0.016 |
| S0 Remission | 48 (53) | 12 (32) | 14 (74) | 22 (67) |  |
| S1 Mild | 17 (19) | 10 (26) | 3 (16) | 4 (12) |  |
| S2 Moderate | 17 (19) | 9 (24) | 2 (11) | 6 (18) |  |
| S3 Severe | 8 (8.9) | 7 (18) | 0 (0) | 1 (3.0) |  |

| Characteristic | Overall N = 252 | Australia N = 90 | Scotland N = 61 | Spain N = 101 | p-value<sup>1</sup> |
| --- | --- | --- | --- | --- | --- |
| **Sampling Characteristics** |  |  |  |  |  |
| Has active symptoms, n (%) | 76 (30) | 29 (32) | 18 (30) | 29 (29) | 0.86 |
| Season, n (%) |  |  |  |  | <0.001 |
| Spring | 152 (60) | 0 (0) | 53 (87) | 99 (98) |  |
| Summer | 0 (0) | 0 (0) | 0 (0) | 0 (0) |  |
| Autumn | 90 (36) | 90 (100) | 0 (0) | 0 (0) |  |
| Winter | 10 (4.0) | 0 (0) | 8 (13) | 2 (2.0) |  |
| **Biochemical Parameters** |  |  |  |  |  |
| Haemoglobin g/L, Median (IQR) | 139 (130 – 148) | 138 (130 – 146) | 135 (127 – 148) | 142 (132 – 151) | 0.024 |
| Red cell count x10<sup>9</sup>/L, Median (IQR) | 4.66 (4.30 – 4.96) | 4.67 (4.30 – 4.87) | 4.54 (4.21 – 4.78) | 4.73 (4.39 – 5.12) | 0.008 |
| White cell count x10<sup>9</sup>/L, Median (IQR) | 7.02 (5.90 – 8.54) | 7.20 (6.00 – 9.00) | 6.90 (6.10 – 7.70) | 6.90 (5.77 – 8.39) | 0.59 |
| Platelets x10<sup>9</sup>/L, Median (IQR) | 270 (231 – 313) | 274 (237 – 323) | 273 (242 – 316) | 264 (220 – 304) | 0.052 |
| Albumin g/L, Median (IQR) | 38 (34 – 42) | 38 (36 – 40) | 39 (38 – 41) | 39 (5 – 45) | 0.47 |
| CRP mg/L, Median (IQR) | 3.0 (1.5 – 5.0) | 2.4 (0.8 – 5.0) | 2.0 (1.0 – 3.5) | 3.0 (3.0 – 5.0) | <0.001 |
| Faecal Calprotectin (µg/g, Median (IQR) | 200 (41 – 477) | 149 (41 – 406) | 93 (36 – 295) | 345 (46 – 1,080) | 0.001 |
| **Drug Therapy at Sampling** |  |  |  |  |  |
| Any steroids, n (%) | 21 (8.3) | 10 (11) | 1 (1.6) | 10 (9.9) | 0.090 |
| Any antibiotics, n (%) | 1 (0.4) | 0 (0) | 0 (0) | 1 (1.0) | >0.99 |
| Any aminosalicylates, n (%) | 65 (26) | 25 (28) | 10 (16) | 30 (30) | 0.15 |
| Azathioprine, n (%) | 32 (13) | 21 (23) | 3 (4.9) | 8 (7.9) | <0.001 |
| Mercaptopurine, n (%) | 6 (2.4) | 5 (5.6) | 0 (0) | 1 (1.0) | 0.057 |
| Methotrexate, n (%) | 2 (0.8) | 1 (1.1) | 1 (1.6) | 0 (0) | 0.52 |
| Infliximab, n (%) | 42 (17) | 25 (28) | 5 (8.2) | 12 (12) | 0.002 |
| Adalimumab, n (%) | 29 (12) | 4 (4.4) | 11 (18) | 14 (14) | 0.023 |
| Vedolizumab, n (%) | 13 (5.2) | 4 (4.4) | 6 (9.8) | 3 (3.0) | 0.17 |
| Ustekinumab, n (%) | 31 (12) | 8 (8.9) | 5 (8.2) | 18 (18) | 0.092 |
| Risankizumab, n (%) | 5 (2.0) | 0 (0) | 2 (3.3) | 3 (3.0) | 0.21 |
| Tofacitinib, n (%) | 1 (0.4) | 0 (0) | 0 (0) | 1 (1.0) | >0.99 |
| Filgotinib, n (%) | 3 (1.2) | 0 (0) | 0 (0) | 3 (3.0) | 0.18 |
| Upadacitinib, n (%) | 9 (3.6) | 4 (4.4) | 3 (4.9) | 2 (2.0) | 0.52 |

<sup>1</sup>Kruskal-Wallis rank sum test; Pearson's Chi-squared test; Fisher's exact test

**Supplementary Table 6.**
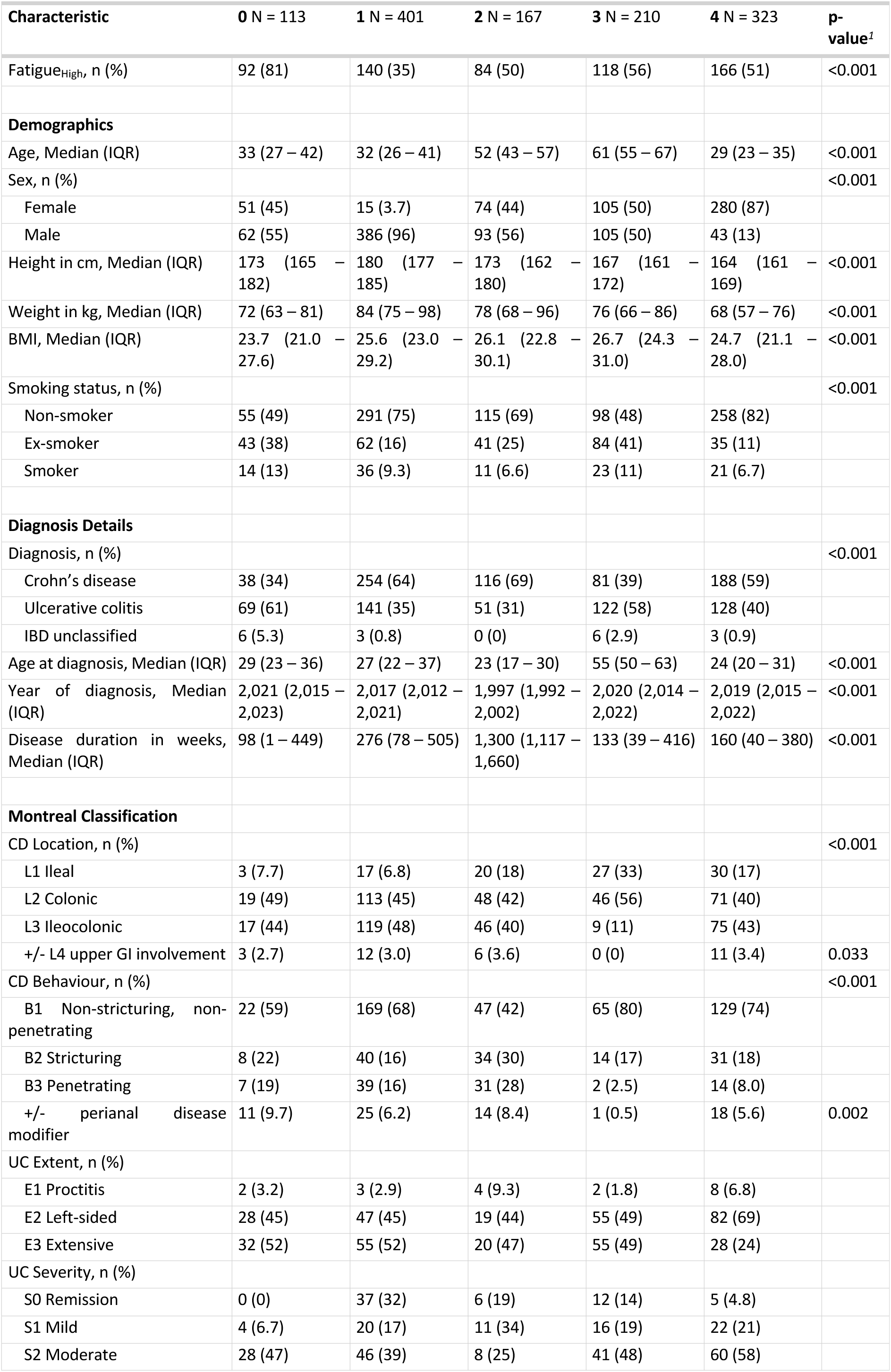
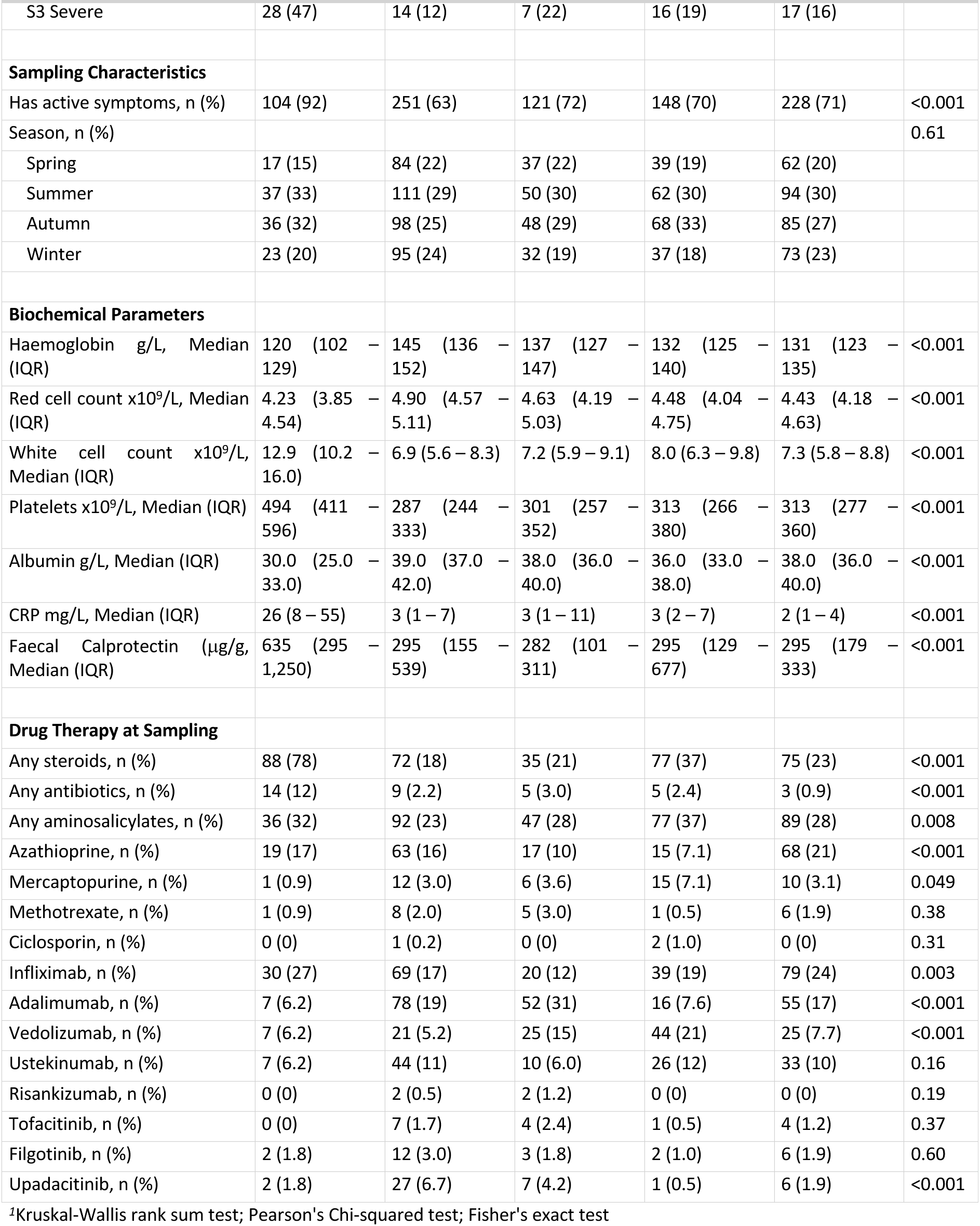
Full Cluster Characteristics.

| Characteristic | 0 N = 113 | 1 N = 401 | 2 N = 167 | 3 N = 210 | 4 N = 323 | p-value<sup>1</sup> |
| --- | --- | --- | --- | --- | --- | --- |
| Fatigue<sub>High</sub>, n (%) | 92 (81) | 140 (35) | 84 (50) | 118 (56) | 166 (51) | <0.001 |
| **Demographics** |  |  |  |  |  |  |
| Age, Median (IQR) | 33 (27 – 42) | 32 (26 – 41) | 52 (43 – 57) | 61 (55 – 67) | 29 (23 – 35) | <0.001 |
| Sex, n (%) |  |  |  |  |  | <0.001 |
| Female | 51 (45) | 15 (3.7) | 74 (44) | 105 (50) | 280 (87) |  |
| Male | 62 (55) | 386 (96) | 93 (56) | 105 (50) | 43 (13) |  |
| Height in cm, Median (IQR) | 173 (165 – 182) | 180 (177 – 185) | 173 (162 – 180) | 167 (161 – 172) | 164 (161 – 169) | <0.001 |
| Weight in kg, Median (IQR) | 72 (63 – 81) | 84 (75 – 98) | 78 (68 – 96) | 76 (66 – 86) | 68 (57 – 76) | <0.001 |
| BMI, Median (IQR) | 23.7 (21.0 – 27.6) | 25.6 (23.0 – 29.2) | 26.1 (22.8 – 30.1) | 26.7 (24.3 – 31.0) | 24.7 (21.1 – 28.0) | <0.001 |
| Smoking status, n (%) |  |  |  |  |  | <0.001 |
| Non-smoker | 55 (49) | 291 (75) | 115 (69) | 98 (48) | 258 (82) |  |
| Ex-smoker | 43 (38) | 62 (16) | 41 (25) | 84 (41) | 35 (11) |  |
| Smoker | 14 (13) | 36 (9.3) | 11 (6.6) | 23 (11) | 21 (6.7) |  |
| **Diagnosis Details** |  |  |  |  |  |  |
| Diagnosis, n (%) |  |  |  |  |  | <0.001 |
| Crohn's disease | 38 (34) | 254 (64) | 116 (69) | 81 (39) | 188 (59) |  |
| Ulcerative colitis | 69 (61) | 141 (35) | 51 (31) | 122 (58) | 128 (40) |  |
| IBD unclassified | 6 (5.3) | 3 (0.8) | 0 (0) | 6 (2.9) | 3 (0.9) |  |
| Age at diagnosis, Median (IQR) | 29 (23 – 36) | 27 (22 – 37) | 23 (17 – 30) | 55 (50 – 63) | 24 (20 – 31) | <0.001 |
| Year of diagnosis, Median (IQR) | 2,021 (2,015 – 2,023) | 2,017 (2,012 – 2,021) | 1,997 (1,992 – 2,002) | 2,020 (2,014 – 2,022) | 2,019 (2,015 – 2,022) | <0.001 |
| Disease duration in weeks, Median (IQR) | 98 (1 – 449) | 276 (78 – 505) | 1,300 (1,117 – 1,660) | 133 (39 – 416) | 160 (40 – 380) | <0.001 |
| **Montreal Classification** |  |  |  |  |  |  |
| CD Location, n (%) |  |  |  |  |  | <0.001 |
| L1 Ileal | 3 (7.7) | 17 (6.8) | 20 (18) | 27 (33) | 30 (17) |  |
| L2 Colonic | 19 (49) | 113 (45) | 48 (42) | 46 (56) | 71 (40) |  |
| L3 Ileocolonic | 17 (44) | 119 (48) | 46 (40) | 9 (11) | 75 (43) |  |
| +/- L4 upper GI involvement | 3 (2.7) | 12 (3.0) | 6 (3.6) | 0 (0) | 11 (3.4) | 0.033 |
| CD Behaviour, n (%) |  |  |  |  |  | <0.001 |
| B1 Non-stricturing, non-penetrating | 22 (59) | 169 (68) | 47 (42) | 65 (80) | 129 (74) |  |
| B2 Stricturing | 8 (22) | 40 (16) | 34 (30) | 14 (17) | 31 (18) |  |
| B3 Penetrating | 7 (19) | 39 (16) | 31 (28) | 2 (2.5) | 14 (8.0) |  |
| +/- perianal disease modifier | 11 (9.7) | 25 (6.2) | 14 (8.4) | 1 (0.5) | 18 (5.6) | 0.002 |
| UC Extent, n (%) |  |  |  |  |  |  |
| E1 Proctitis | 2 (3.2) | 3 (2.9) | 4 (9.3) | 2 (1.8) | 8 (6.8) |  |
| E2 Left-sided | 28 (45) | 47 (45) | 19 (44) | 55 (49) | 82 (69) |  |
| E3 Extensive | 32 (52) | 55 (52) | 20 (47) | 55 (49) | 28 (24) |  |
| UC Severity, n (%) |  |  |  |  |  |  |
| S0 Remission | 0 (0) | 37 (32) | 6 (19) | 12 (14) | 5 (4.8) |  |
| S1 Mild | 4 (6.7) | 20 (17) | 11 (34) | 16 (19) | 22 (21) |  |
| S2 Moderate | 28 (47) | 46 (39) | 8 (25) | 41 (48) | 60 (58) |  |
| S3 Severe | 28 (47) | 14 (12) | 7 (22) | 16 (19) | 17 (16) |  |
| **Sampling Characteristics** |  |  |  |  |  |  |
| Has active symptoms, n (%) | 104 (92) | 251 (63) | 121 (72) | 148 (70) | 228 (71) | <0.001 |
| Season, n (%) |  |  |  |  |  | 0.61 |
| Spring | 17 (15) | 84 (22) | 37 (22) | 39 (19) | 62 (20) |  |
| Summer | 37 (33) | 111 (29) | 50 (30) | 62 (30) | 94 (30) |  |
| Autumn | 36 (32) | 98 (25) | 48 (29) | 68 (33) | 85 (27) |  |
| Winter | 23 (20) | 95 (24) | 32 (19) | 37 (18) | 73 (23) |  |
| **Biochemical Parameters** |  |  |  |  |  |  |
| Haemoglobin g/L, Median (IQR) | 120 (102 – 129) | 145 (136 – 152) | 137 (127 – 147) | 132 (125 – 140) | 131 (123 – 135) | <0.001 |
| Red cell count x10<sup>9</sup>/L, Median (IQR) | 4.23 (3.85 – 4.54) | 4.90 (4.57 – 5.11) | 4.63 (4.19 – 5.03) | 4.48 (4.04 – 4.75) | 4.43 (4.18 – 4.63) | <0.001 |
| White cell count x10<sup>9</sup>/L, Median (IQR) | 12.9 (10.2 – 16.0) | 6.9 (5.6 – 8.3) | 7.2 (5.9 – 9.1) | 8.0 (6.3 – 9.8) | 7.3 (5.8 – 8.8) | <0.001 |
| Platelets x10<sup>9</sup>/L, Median (IQR) | 494 (411 – 596) | 287 (244 – 333) | 301 (257 – 352) | 313 (266 – 380) | 313 (277 – 360) | <0.001 |
| Albumin g/L, Median (IQR) | 30.0 (25.0 – 33.0) | 39.0 (37.0 – 42.0) | 38.0 (36.0 – 40.0) | 36.0 (33.0 – 38.0) | 38.0 (36.0 – 40.0) | <0.001 |
| CRP mg/L, Median (IQR) | 26 (8 – 55) | 3 (1 – 7) | 3 (1 – 11) | 3 (2 – 7) | 2 (1 – 4) | <0.001 |
| Faecal Calprotectin (µg/g, Median (IQR) | 635 (295 – 1,250) | 295 (155 – 539) | 282 (101 – 311) | 295 (129 – 677) | 295 (179 – 333) | <0.001 |
| **Drug Therapy at Sampling** |  |  |  |  |  |  |
| Any steroids, n (%) | 88 (78) | 72 (18) | 35 (21) | 77 (37) | 75 (23) | <0.001 |
| Any antibiotics, n (%) | 14 (12) | 9 (2.2) | 5 (3.0) | 5 (2.4) | 3 (0.9) | <0.001 |
| Any aminosalicylates, n (%) | 36 (32) | 92 (23) | 47 (28) | 77 (37) | 89 (28) | 0.008 |
| Azathioprine, n (%) | 19 (17) | 63 (16) | 17 (10) | 15 (7.1) | 68 (21) | <0.001 |
| Mercaptopurine, n (%) | 1 (0.9) | 12 (3.0) | 6 (3.6) | 15 (7.1) | 10 (3.1) | 0.049 |
| Methotrexate, n (%) | 1 (0.9) | 8 (2.0) | 5 (3.0) | 1 (0.5) | 6 (1.9) | 0.38 |
| Ciclosporin, n (%) | 0 (0) | 1 (0.2) | 0 (0) | 2 (1.0) | 0 (0) | 0.31 |
| Infliximab, n (%) | 30 (27) | 69 (17) | 20 (12) | 39 (19) | 79 (24) | 0.003 |
| Adalimumab, n (%) | 7 (6.2) | 78 (19) | 52 (31) | 16 (7.6) | 55 (17) | <0.001 |
| Vedolizumab, n (%) | 7 (6.2) | 21 (5.2) | 25 (15) | 44 (21) | 25 (7.7) | <0.001 |
| Ustekinumab, n (%) | 7 (6.2) | 44 (11) | 10 (6.0) | 26 (12) | 33 (10) | 0.16 |
| Risankizumab, n (%) | 0 (0) | 2 (0.5) | 2 (1.2) | 0 (0) | 0 (0) | 0.19 |
| Tofacitinib, n (%) | 0 (0) | 7 (1.7) | 4 (2.4) | 1 (0.5) | 4 (1.2) | 0.37 |
| Filgotinib, n (%) | 2 (1.8) | 12 (3.0) | 3 (1.8) | 2 (1.0) | 6 (1.9) | 0.60 |
| Upadacitinib, n (%) | 2 (1.8) | 27 (6.7) | 7 (4.2) | 1 (0.5) | 6 (1.9) | <0.001 |

<sup>1</sup>Kruskal-Wallis rank sum test; Pearson's Chi-squared test; Fisher's exact test

**Supplementary Figure 1.**
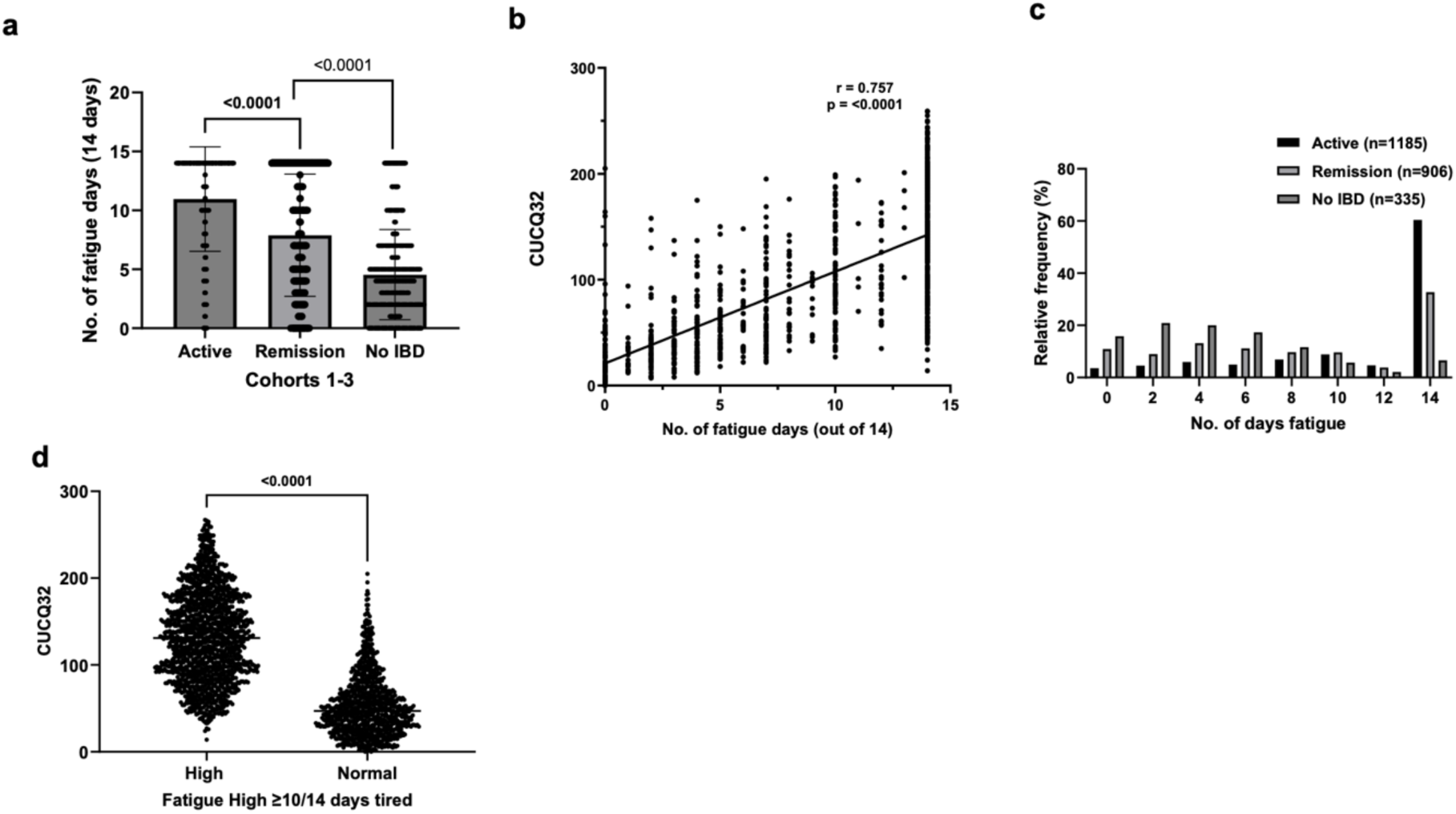
Fatigue days differ by disease status and correlate with CUCQ32 scores. (a) Median number of fatigue days differed significantly between patients with active I8D, patients in remission, and non-I8D controls (all cohorts combined). (b) Fatigue days reported in the CUCQ fatigue question correlated significantly with overall CUCQ32 scores. (c) Reported fatigue days were non-normally distributed, with a median of 10 used to classify fatigue as Fatigue_High_ or Fatigue_Low_ (d) Overall CUCQ32 scores were significantly higher in the Fatigue-high group compared to the Fatigue-low group.

**Supplementary Figure 2.**
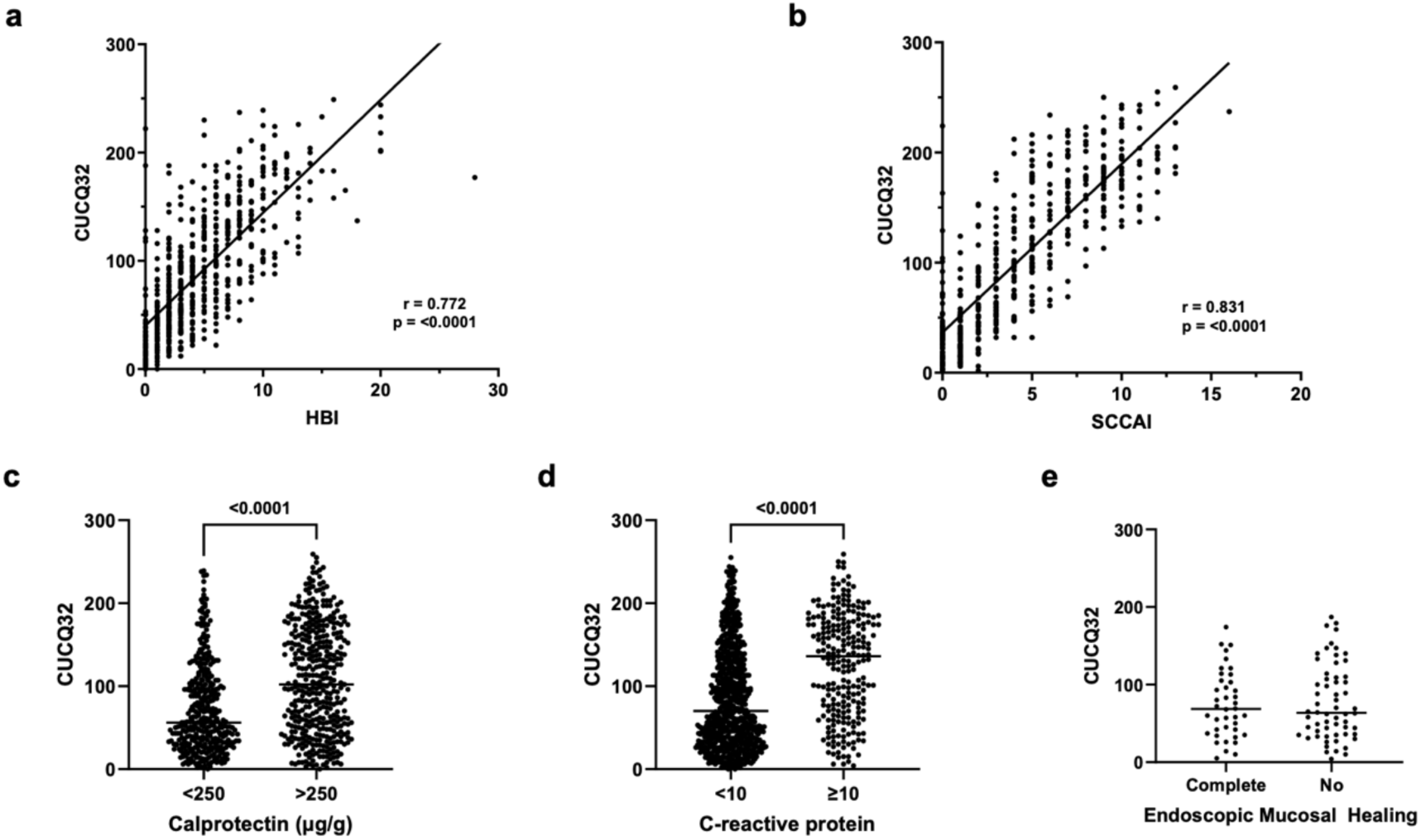
CUCQ32 scores correlate with clinical activity scores and inflammatory biomarkers but not with endoscopic healing. (a) CUCQ32 scores are highly correlated with Harvey-Bradshaw Index scores in Crohn’s disease. (b) CUCQ32 scores are highly correlated with Simple Clinical Colitis Activity Index scores in ulcerative colitis. (c) CUCQ32 scores are significantly higher in patients with elevated faecal calprotectin (>250 µg/g). (d) CUCQ32 scores are significantly higher in patients with elevated C-reactive protein (>10 mg/L). (e) No significant difference in CUCQ32 scores was observed between patients with complete mucosa! healing and those without, in Cohort 3 (MUSIC).

**Supplementary Figure 3.**
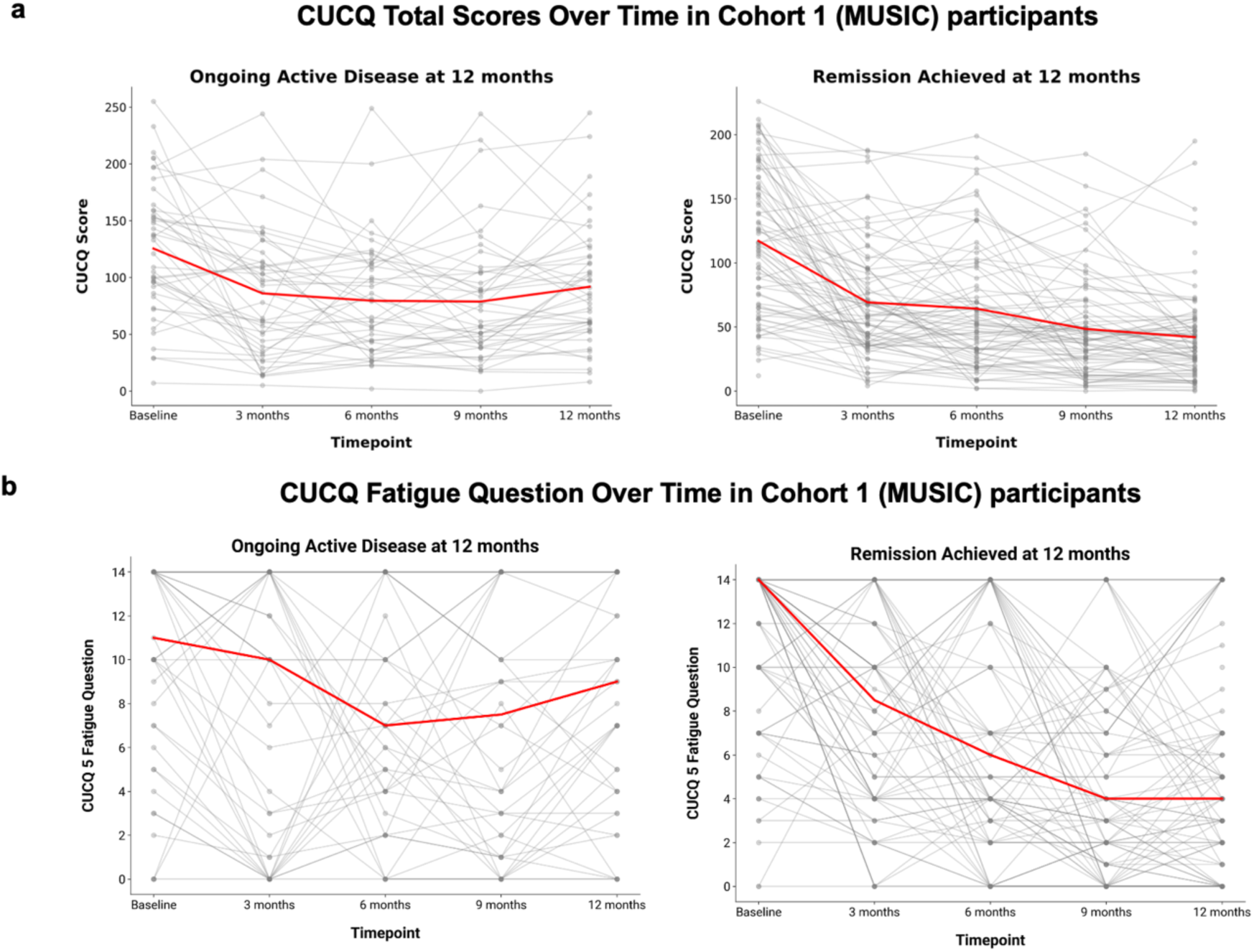
CUCQ Longitudinal Characteristics. (a) & {b) Evolution of CUCQ32 total scores (top) and CUCQ32 fatigue question scores {bottom) over time in patients with ongoing disease activity versus remission in the MUSIC cohort. Red lines represent the mean for CUCQ32 total scores and the median for the fatigue question scores (Cohort 1).

**Supplementary Figure 4.**
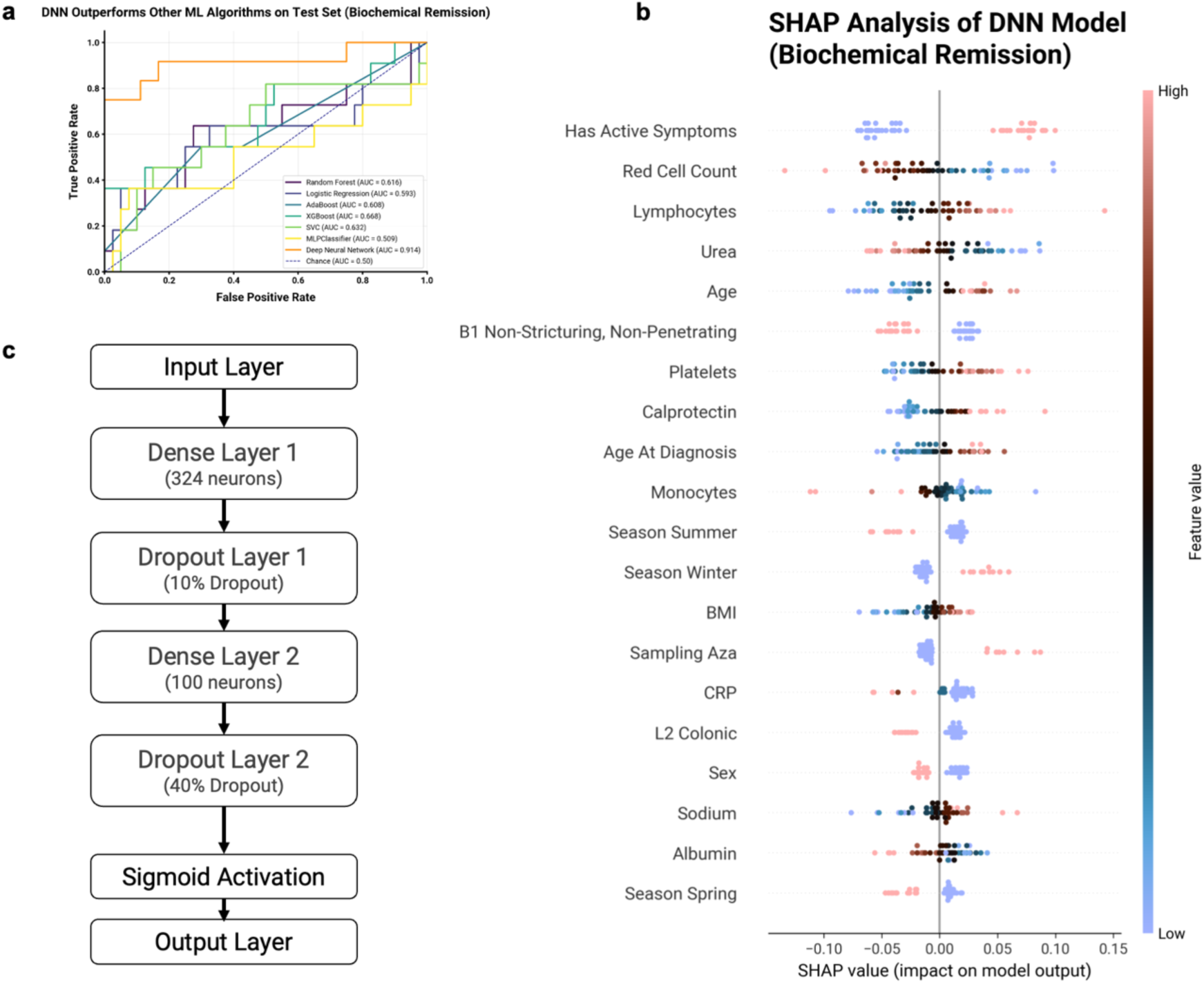
Model performance, features associated with fatigue prediction in the biochemical remission cohort and final DNN architecture. (a) Model performance (ALIC) in the biochemical remission cohort. The dataset size for this cohort is smaller, and predicting fatigue is more challenging. (b) SHAP summary plot for the deep neural network (DNN) in the biochemical remission cohort. The top features associated with fatigue include active IBD symptoms, low red blood cell counts, high lymphocyte counts, low urea levels, and older age. Seasonality factors (summer and winter) have smaller contributions but are captured within the model. (c) Schematic overview of final deep neural network architecture.

